# Evidence And Mechanisms For Embolic Stroke In Contralateral Hemispheres From Carotid Artery Sources

**DOI:** 10.1101/2023.04.20.23288892

**Authors:** Ricardo Roopnarinesingh, Michelle Leppert, Debanjan Mukherjee

## Abstract

Disambiguation of embolus etiology in embolic strokes is often a clinical challenge. One common source of embolic stroke is the carotid arteries, with emboli originating due to plaque build up, or perioperatively during revascularization procedures. While it is commonly thought that thromboemboli from carotid sources travel to cerebral arteries ipsilaterally, there are existing reports of contralateral embolic events which complicate embolus source destination relationship for carotid sources. Here, we hypothesize that emboli from carotid sources can travel to contralateral hemispheres, and that embolus interactions with collateral hemodynamics in the Circle of Willis influences this process. We use a patient-specific computational embolus-hemodynamics interaction model developed in prior works to conduct an *in silico* experiment spanning 4 patient vascular models, 6 Circle of Willis anastomosis variants, and 3 different thromboembolus sizes released from left and right carotid artery sites. This led to a total of 144 different experiments, estimating trajectories and distribution of approximately 1.728 million embolus samples. Across all cases considered, emboli from left and right carotid sources showed non-zero contralateral transport (p value *<* 0.05). Contralateral movement revealed a size-dependence, with smaller emboli traveling more contralaterally. Detailed analysis of embolus dynamics revealed that collateral flow routes in Circle of Willis played a role in routing emboli, and transhemispheric movement occurred through the anterior and posterior communicating arteries in the Circle of Willis. We generated quantitative data demonstrating the complex dynamics of finite size thromboembolus particles as they interact with pulsatile arterial hemodynamics, and traverse the vascular network of the Circle of Willis. This leads to a non-intuitive source-destination relationship for emboli originating from carotid artery sites, and emboli from carotid sources can potentially travel to cerebral arteries on contralateral hemispheres.

## 1 Introduction

Stroke remains a leading a cause of death and disability worldwide, with a significant fraction of cases being acute ischemic strokes of embolic origin. However, these embolus sources are not always known. Embolic Strokes of Undetermined Sources (ESUS) have received increasing attention in recent years [1, 2]. ESUS cases refer to an occlusion in the cerebral artery by an embolus which could have originated from multiple potential co-occurring sources, and thereby their etiology is challenging to disambiguate. While it is reckoned that approximately 25-30% of ischemic strokes can be categorized as ESUS [3], some studies have reported that up to 60% of ischemic strokes in some cohorts, especially young adults, can be categorized as ESUS [4]. This motivates the need to improve clinical identification of the embolus source. Improved methods of identifying embolus source, and source-destination relationship would allow for increased treatment efficacy and reduced recurrent events with treatment based on the etiology [2]. This source-destination relationship for emboli is determined by an interplay of embolus properties, patient vascular anatomy for the heart-brain pathway, and patient hemodynamics. Specifically, the carotid arteries are a critical source of stroke due to their susceptibility to the buildup of atherosclerotic plaques [5, 6]. It is widely regarded that the most relevant cause of strokes from carotid artery diseases are thromboemboli [6]. Rupture of unstable plaques, and associated thromboembolic events, can lead to embolic debris to travel into the Circle of Willis (CoW) and cause an embolic stroke. This can also occur peri-operatively during carotid revascularization therapies such as carotid artery stenting (CAS) or carotid endarterectomy (CAE) [7, 8]. The common notion about carotid embolus source-destination relationship is that embolus from the carotid arteries travel ipsilaterally to the cerebral arteries or to the same hemisphere relative to the carotid side. Although this is true for a large proportion of cases, there have been many reported instances of emboli traveling contralaterally, or transhemispheric passage of emboli, from the carotid arteries where the embolus intermixes or crosses between the brain hemispheres relative to the source carotid [9–12]. Specifically, despite contralateral stroke events being less common, they can be a key contributor to the difficulty in disambiguating stroke etiology as these cases can be the non-decipherable occurrences comprising ESUS patients. It remains challenging to study how emboli released from left/right carotid arteries pass transhemispherically using traditional imaging and animal model studies, and modern *in silico* techniques can prove to be a viable alternative. In a series of prior works, we have established a patient-specific *in silico* flow-embolus interaction model that enables us to quantitatively study the source-destination relationship for the transport of emboli to the brain [13–16]. We have used this model to demonstrated the transport and distribution of cardiogenic emboli and emboli from aortic arch, and illustrate the dependency of embolus distribution on embolus properties, local flow features, and arterial network anatomy. Here, we use our model to conduct a parametric *in silico* study to understand how emboli from carotid artery sites travel to the six cerebral vessels of the CoW as a function of size, CoW anatomy, and laterality of carotid release sites.

## 2 Methods

### 2.1 Image-Based Modeling of Vascular Anatomy

The CoW anatomy has significant variations in topology and anastomosis across subjects, commonly with one or more communicating vessels missing or hypoplastic [16–19]. Using a meta-analysis of literature on reported CoW anatomical variations, and a ranking statistics approach outlined in our prior work [16], 5 most frequent variations of the CoW anatomy were considered for this study. These variations involved an absent anterior communicating artery (*labelled as AcoA for this study*), absent left and right P1 connectors (*labelled as LP1 and RP1 for this study*), and absent left and right communicating arteries (*labelled as L*.*Comm and R*.*Comm for this study*). Together with the complete CoW, this led to a total of 6 CoW anatomical variants considered in this study. Fig 1 shows the differences between these CoW anatomies with reference to corresponding missing vessels along with labels for various cerebral arteries. These labels and identifiers are used throughout the paper. Four patient anatomical geometries, used in our previous works [14, 16], were selected from a set of computed tomography (CT) images from an Institutional Review Board (IRB) approved Screening Technology and Outcome Project in Stroke (STOP-Stroke) database [20]. The STOP-Stroke database comprised 675 patients who were diagnosed with a Stroke or a Transient Ischemic Attack (TIA), categorized into Large Vessel Occlusion (LVO) and non-LVO cases, with 46% of the stroke cases identified as LVO. For this study, we utilized the non-contrast and contrast-enhanced CT images for retrospective, secondary, computational analysis purposes; and rest of the data from the database was not utilized. Consequently, no additional IRB approval was required for this study. These patients were selected from the overall database to ensure that: (a) they had a complete CoW anatomy; and (b) the branching vessels at the aortic branch were segregated such that there was no fused vessels or bovine arch anatomy. For each patient, artery lumen segmentations were generated from the contrast-enhanced CT images using a 2D planar segmentation approach implemented in the open-source cardiovascular modeling package SimVascular [21]. For each patient segmentation, the cerebral arteries up until the M1, A1, and P1 segments of the MCA, ACA, and PCA respectively, were included. These artery segmentations were then lofted to generate a 3D solid model of the complete arterial pathway from the aorta to the CoW. Subsequently, the CoW variation models were created by removing the respective absent CoW artery paths and segments from the complete patient CoW anatomy segmentation, and subsequently re-lofting the new geometry to produce the 3D model of the corresponding incomplete CoW anastomosis. Details of this workflow are outlined in prior work [16]. By removing the arteries from a single patient with a complete CoW anatomy instead of introducing another patient with that specific CoW variation, we eliminate the multiple possible interpatient variabilities in our *in silico* investigation. Using four patients each with six CoW anastomoses, 24 total patient CoW vascular models were generated. Figure 1 shows each patient heart-to-brain pathway along with diagrams that correspond to the CoW anastomosis variation.

**Figure 1:**
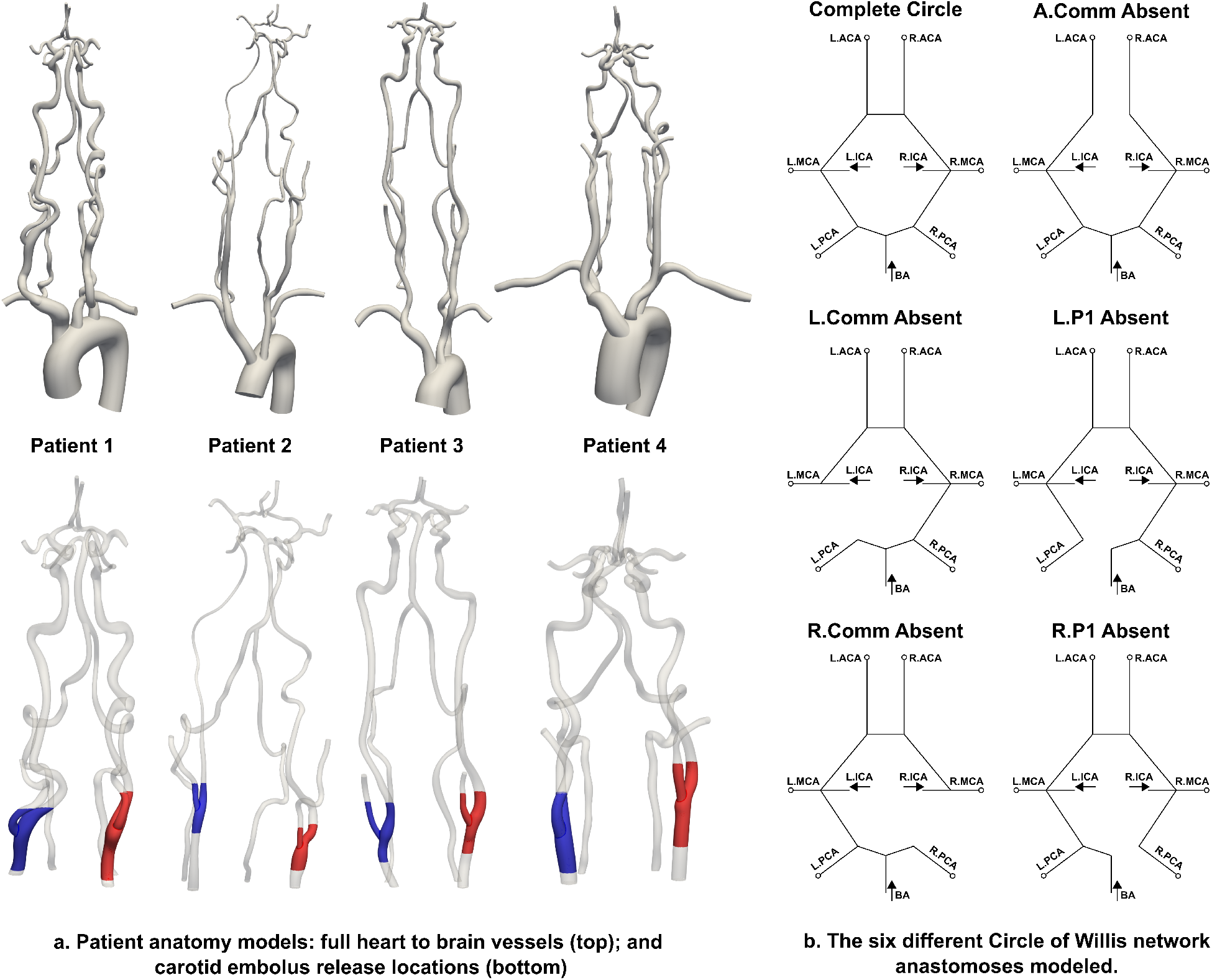
Complete heart-to-brain pathway was modeled for four patients combining 6 Circle of Willis (CoW) anatomical variations (a. top). Emboli were released from both carotid arteries for all four patients (a. bottom). The six variations of the CoW of listed are shown in b with each absent artery being depicted relative to the complete CoW anastomosis.

### 2.2 Hemodynamics Simulation

Three-dimensional simulations of blood flow using computational fluid dynamics (CFD) was conducted for each of the 24 models. For these simulations, each model was discretized into a computational mesh comprising approximately 6.0 to 6.5 million linear tetrahedral elements with the average element size being between 0.36 and 0.52 mm for each model. These parameters were chosen through mesh refinement analysis described in prior works [16]. Meshes were generated using the TetGen software tool, integrated within the SimVascular package. Blood flow was modeled using the classical Navier-Stokes equations for momentum balance and the continuity equation for mass balance. A Petrov-Galerkin stabilized finite-element method built into the SimVascular package was used to solve the mass and momentum equations for the velocity and pressure fields [22, 23]. A summary of the underlying mathematical equations is presented in the Supplementary Material. A pulsatile flow velocity profile was designated as an inlet boundary condition at the aortic root of the model. The profile was derived based on magnetic resonance measurements reported in [24]. This specified inlet flow led to a consistent average volumetric flow rate of 79 mL/s as the cardiac output (CO) for all models. To account for influence of the downstream vascular beds at each of the truncated arterial outlets, flow resistance boundary conditions were assigned to each outlet of the model. An average systolic and diastolic blood pressure of 120 and 80 mmHg was assumed, leading to a mean arterial pressure (MAP) of 93.33 mmHg, which was used to compute a total arterial resistance (TAR) as MAP/CO. Subsequently, a proportion of the TAR was assigned to each vessel outlet based on target flow divisions as outlined in prior work [14]. Briefly: (a) 65% of total flow was assumed to exit the descending aorta [25]; (b) flow rates at the six cerebral artery outlets were assigned based on measured MR data reported in [26]; and (c) the remainder volumetric flow was designated to exit the external carotid and the subclavian artery outlets based on their cross-sectional area [27]. Multiple steady-state flow simulations were conducted to tune the outlet resistance values for the boundary conditions, to match the target flow rates described here [14, 15]. Once obtained, the CO, MAP, and outlet resistance values were kept the same across all patients and CoW variations to allow for controlled simulations of embolus transport. A total of three cardiac cycles of blood flow were simulated for each model, and the results from the third cardiac cycle was used further for embolus transport calculations. Extensive details on each underlying methodological piece have been described in a series of prior works [15, 16].

### 2.3 Embolus Transport

Transport of thrombo-emboli across the arteries was modeled using a custom modified version of the Maxey-Riley equation [15, 28], which calculates the trajectories of individual particles moving through a fluid flow based on local velocities and particle properties. Individual thrombo-emboli were modeled as idealized spherical particles in accordance with the underlying physics of the Maxey-Riley equation, with a density of 1.1 g/cm^3^ and elasticity of 1667.64 Pa as obtained from literature [29, 30]. Our modifications to the classical form of Maxey-Riley equations included incorporating: (a) lift force produced by near-wall shear gradients; (b) collisions between particles and the artery wall; and (c) elastohydrodynamic lubrication effects near-wall. These are in addition to standard drag forces, added mass effects, and forces from the undisturbed background flow. Mathematical details of each of these physical forces are provided in the Supplementary Material. Particle-particle collisions were not taken into account as the trajectory of individual particles were modeled independent of each other. Flow velocity data from the third cardiac cycle of hemodynamics simulations was stitched together 5 times to create a representative flow-field spanning 5 cardiac cycles that has no cycle-to-cycle flow variations. This was used to compute the embolus trajectories across a 5 cardiac cycle duration. The integration of each particle trajectory was completed using an Euler time integration method with a time-step of 0.05 ms. The embolus computation workflow is implemented in a set of custom computer codes built using C and Python with the Visualization Toolkit Libraries (VTK) for pre-processing and post-processing of flow velocity data and particle data respectively. Extensive details on the individual mathematical expressions and algorithms can be found in our prior works [14–16].

### 2.4 In Silico Experiment Design

For this study, we used the simulation tools described in Section 2.2 and 2.3 to set up a parametric set of numerical experiments for embolus transport originating from carotid artery sources. The outline of the experiment design is illustrated in Figure 2. Specifically, for each of the 24 patient CoW anatomical variants modeled as stated in Section 2.1, representative thrombo-embolic particles were released along the walls of the left and the right carotid arteries as indicated in Figure 1. Approximately 12,000 embolic particle samples were released for each case. Three different embolus sizes were used: 500 microns, 700 microns, and 1000 microns. As stated above, each embolic particle trajectory integration was completed for 5 cardiac cycles during which all 12,000 particles exited into one of the vessel outlets. Across all parameter combinations, this led to a total of 144 simulations, each comprising 12,000 embolic particle samples, leading to distribution data spanning a total 1.728 million embolus samples. From each of the 144 simulations, we obtained the embolus distribution to the 6 cerebral arteries (left and right M1, A1, and P1) as a number fraction defined as N_*k*_/N_*cow*_ where N_*k*_ is the number of emboli that reached a specific cerebral artery (identified by *k*), and N_*cow*_ is the total number of emboli that reached the CoW from the carotid source.

**Figure 2:**
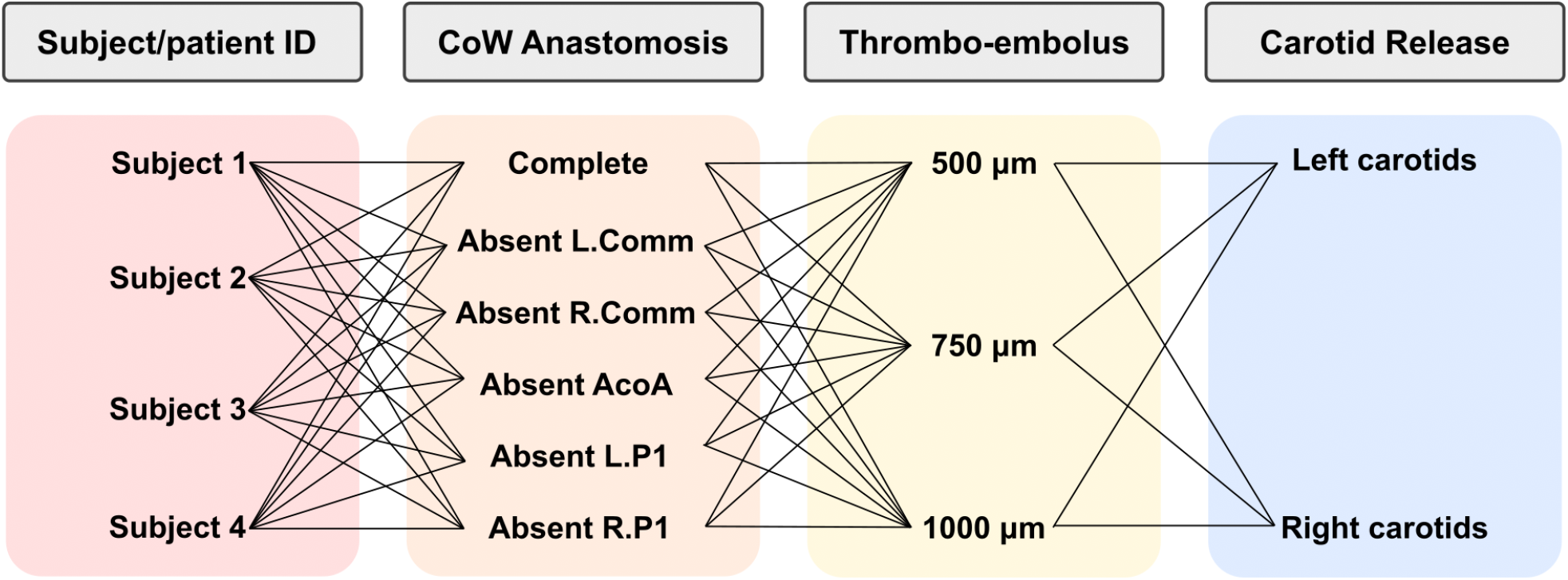
The structure of in silico experiments comprising a total of 144 different embolus transport simulations. These parameters include four patients, six CoW variations, three embolus sizes, and two release locations.

## 3 Results

### 3.1 Total Embolus Distribution

The total combined distribution statistics of emboli to each cerebral arteries across all the simulation cases are presented in Figure 3, categorized based on: (a) left vs right carotid release sites; and (b) embolus size. The first sample box plot in each panel represents the flow distribution to the respective cerebral artery across all simulation models. We observe that the majority of emboli travel to ipsilateral hemisphere sites: that is, majority of emboli reach cerebral arteries on the same hemisphere side as the respective carotid release. We also observe that the majority of the emboli distribute to the left and right M1 segment outlets. Both of these observations are commonly expected based on the natural direct anatomical connection of the ICA into the MCA for left and right. However, the data in Figure 3 also indicate that there is a finite non-zero proportion of emboli that move into cerebral arteries on the contralateral side: that is, emboli reach cerebral arteries on the opposite hemisphere side as the respective carotid release. Based on our *in silico* experiment results, the contralateral distribution fractions are more notable in the ACA and MCA segments.

**Figure 3:**
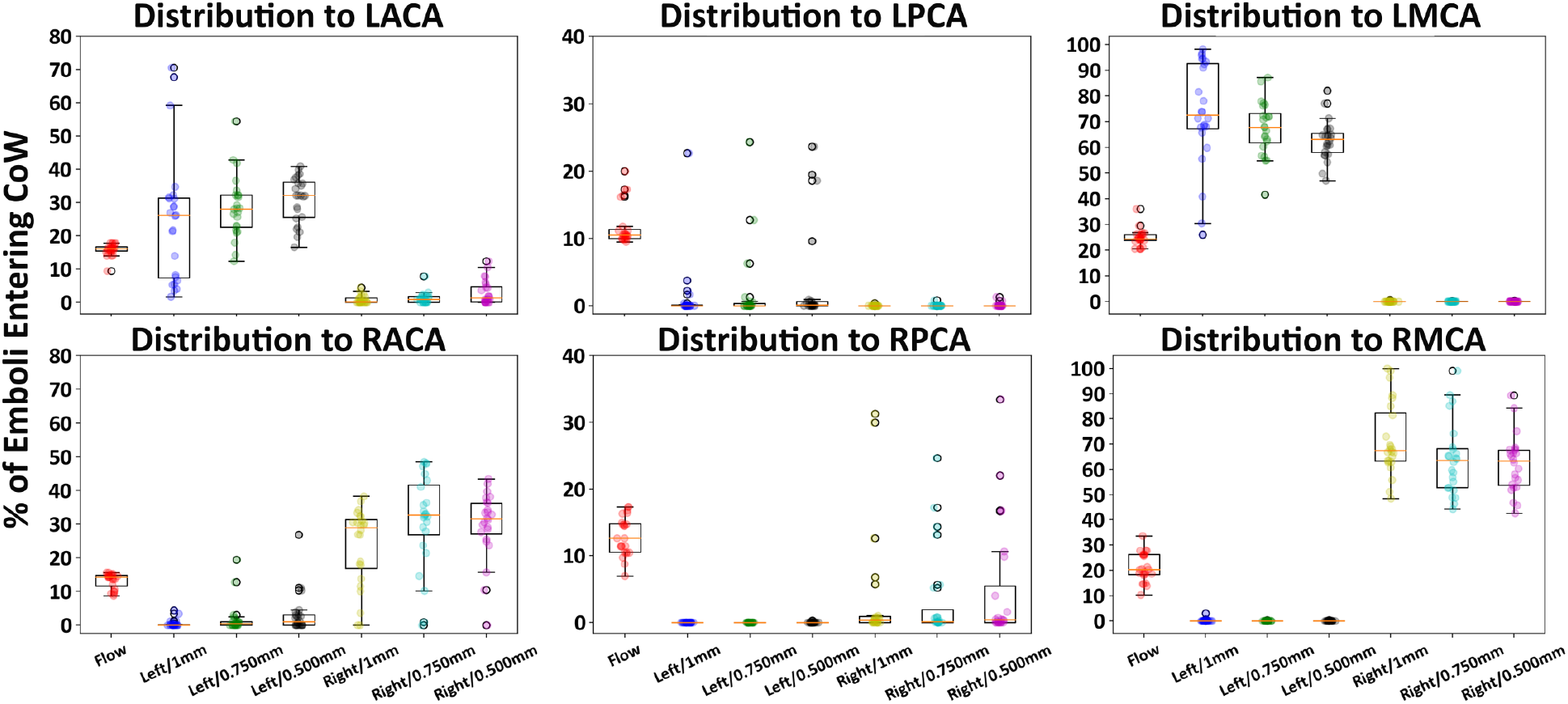
Total distribution of emboli to each cerebral vessel across all simulations. Flow distribution to each cerebral vessel is depicted using the tag ‘Flow’. The distribution is represented as percentage of emboli/flow that reached the specific vessel to the total amount of emboli/flow entering the Circle of Willis.

Throughout all of the cases considered, we note that embolus distribution to the cerebral arteries notably differs from flow distribution, thereby indicating that flow distribution alone is not a suitable indicator of the fractions in which emboli originating from a specific source will reach a cerebral artery. Specifically, a one-sample non-parametric Wilcoxon test for difference between the embolus distribution across all sizes and the flow distribution to each of the 6 cerebral arteries was conducted. The tests led to a p-value *<*= 0.05 for RMCA, LMCA, RPCA, and LPCA respectively; while p-values for LACA and RACA were 0.06 and 0.32 respectively. This distinction from flow distribution has been also discussed in several prior works [13, 16, 31]; and considering the CoW anastomosis variations considered here, the results indicate that the proximal collateral design of the CoW along with embolus size-dependent transport causes substantial variabilities in embolus destination and distribution. The variabilities were statistically characterized across all *in silico* experiments using the coefficient of variation of embolus distribution fractions to each of the 6 cerebral arteries. The coefficient of variation was calculated as the ratio of sample standard deviation and sample mean for the embolus distribution fraction to each cerebral artery across all models and embolus sizes. The estimated coefficients of variation were 1.09, 1.04, and 3.87 respectively for LACA, LMCA, and LPCA; and 1.11, 1.05, and 3.06 respectively for RACA, RMCA, RPCA - indicating high variability with all values being greater than 1.0, and most variability observed in RPCA and LPCA embolus distributions for emboli originating from carotid artery sources.

### 3.2 Extent of Contralateral Movement

In Figure 4, we specifically illustrate the extent of contralateral movement of emboli originating from the left and right carotid arteries, categorized across the 6 different CoW anastomoses and across the 4 patients. Specifically, we depict the percentage of emboli entering the CoW that travel to a cerebral artery contralateral to the carotid site it was released from. The last bar in Figure 4 for each CoW anatomical variant depicts the average of the contralateral embolus distribution across all 4 patients. As stated in Section 3.1, contralateral movement of emboli is observed in the majority of models. Specifically, for both left carotid and right carotid releases, 18 out of the 24 patient CoW anatomical variant models considered led to non-zero contralateral embolus distribution. Statistically, the null hypothesis that emboli released from left as well as right carotid sites do not travel contralaterally (that is the proportion of contralaterally traveling emboli = 0) was rejected with p-values of 0.0003 and 0.0001 respectively (based on a one-sample non-parametric Wilcoxon rank test). For emboli originating from the left carotid sites, CoW variants with missing RComm and missing RP1 led to the highest percentage of contralateral embolus distribution. For emboli originating from the right carotid sites, CoW variant with missing LP1 led to the highest perventage of contralateral embolus distribution. Additionally, cases with zero contralateral embolus distribution most consistently involved the CoW anastomosis with the missing AcoA, with only one case with missing AcoA leading to contralateral embolus transport.

**Figure 4:**
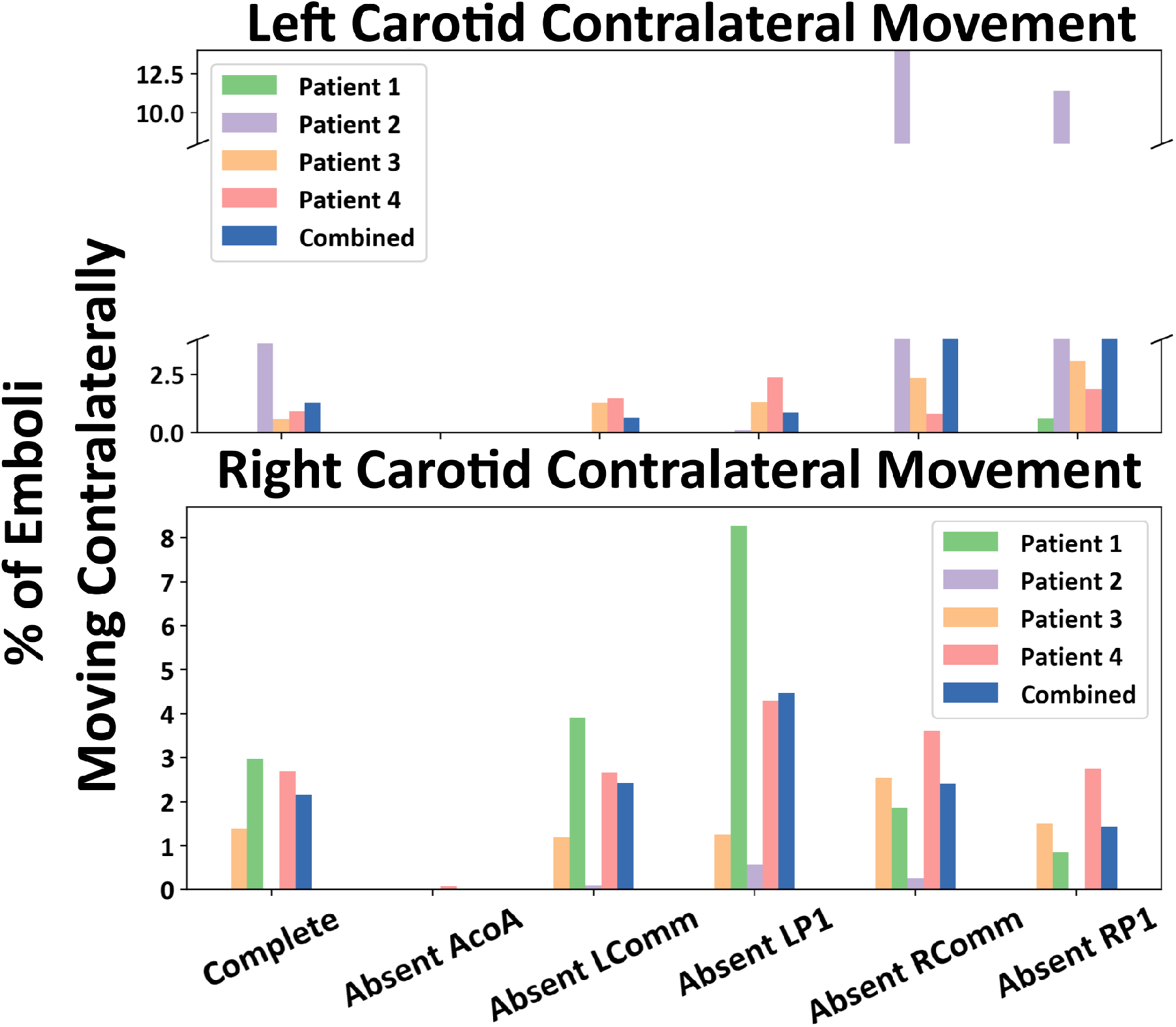
The proportion of contralateral movement for both the left and right carotid releases across all six CoW variations. Left figure shows the percentage of emboli entering the CoW that are released from the left carotid and travel to the right cerebral arteries. The right figure shows the percentage of emboli entering the CoW that are released from the right carotid and travel to the left cerebral arteries.

### 3.3 Embolus Size Dependence

Figure 5 illustrates the dependency of contralateral movement on embolus size. Panels a., b., and c. respectively depict the percentage of contralaterally traveling emboli that are 500 *μ*m, 750 *μ*m, and 1000 *μ*m diameter. Similar to Figure 4, each panel depicts the distribution categorized across the 6 CoW anatomical variants for each of the 4 patient anatomical models considered. For each CoW anatomical variant, the 5’th bar depicts the average of the contralateral embolus distribution across all 4 patients. The results show that in the simulation cases considered, smaller emboli have a greater tendency to distribute contralaterally, with the 1000 micron embolus having the smallest proportion of contralateral embolus and the 500 micron embolus having the largest proportion of contralaterally traveling emboli.

**Figure 5:**
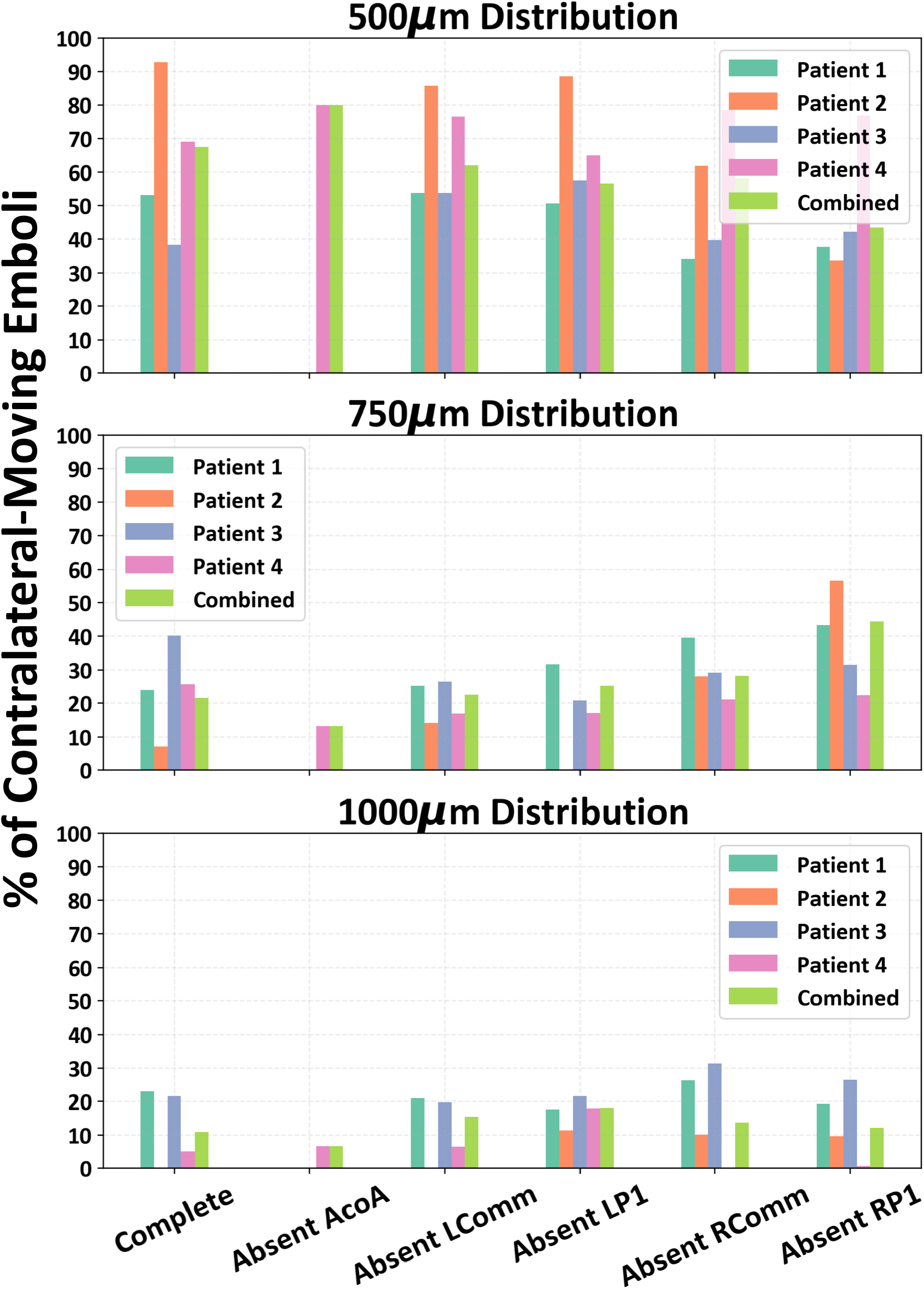
The percentage of contralateral embolus movement decomposed based on embolus size. The illustration indcates that smaller emboli held a larger proportion of contralateral movement in comparison to larger.

## 4 Discussion

Simulation cases considered in this study demonstrate contralateral movement of emboli originating from carotid artery sites leading to otherwise non-intuitive embolus distributions. We note that CoW anastomosis is a key factor influencing the extent of contralateral embolus movement. In a prior work, we have quantified how the communicating arteries of the CoW recruit and route flow and cardiogenic emboli into the 6 major cerebral arteries across different CoW anastomosis [16]. As an embolus enters the CoW from the three major inlet vessels (left and right ICA and Basilar artery), it can get entrained in the flow patterns across the CoW communicating vessels, and ultimately travel across more than one vessel segments to reach its destination M1/A1/P1 segment in the left/right hemisphere. This interaction of collateral flow in CoW with the embolus is therefore one key factor that determines the propensity of contralateral embolus distribution. Specifically, emboli entering the CoW from one of the carotid arteries directly feeding the CoW may not reach a cerebral artery on the same side based on anatomical considerations alone, but instead be entrained in CoW collateral flow to traverse several vessel segments and reach a cerebral artery on the opposite hemisphere.

As also observed in Section 3.3, contralateral embolus distribution is influenced by embolus size. This can also be interpreted in terms of embolus interaction with collateral flow. Specifically, we refer to a physical parameter named momentum response time (MRT), which is a surrogate descriptor for the time that a particle takes to respond to changes in the background fluid flow that it is immersed in. Additional mathematical definitions and details are provided in Supplementary Material. For the same material (say, thrombo-embolus considered here), the MRT is proportional to the square of the particle diameter. Thus, a smaller particle will have lesser MRT, and will respond to and get entrained by the background flow faster. Particle size is directly related to particle inertia, and particles with finite inertia will therefore have a non-zero MRT and will not exactly follow the path of the flow. These factors taken together explain the observations that embolus distribution differs from flow distribution, and the distribution ultimately has a dependency on size. Specifically, the smaller emboli may be recruited by the collateral flow pathways of the CoW anastomosis more rapidly than the larger emboli, thereby explaining why smaller emboli show greater extent of contralateral movement aided by flow routing across the CoW vessels. This aspect of flow-embolus interaction physics has also been discussed in mathematical detail in conjunction with embolus material properties and ratio of material density between embolus and blood in our prior work [15, 16]

Amongst the communicating vessels in the CoW, the Anterior Communicating Artery (AcoA) has been known to play a key role in collateralization of flow across hemispheres. High patency of AcoA is often related to contralateral microemboli as well [32]. Here, detailed quantitative data from our *in silico* experiments further support and substantiate this idea. We have shown in Figure 4 that most cases with 0 contralateral embolus distribution correspond to CoW anastomosis with a missing AcoA. Additionally, we present a case study of Patient 1 in Figure 6, panel a., where left posterior communicating artery is missing and we observe the collateral flow influencing the contralateral susceptibility for both the left and right carotid releases. This patient had 8% of emboli travel contralaterally, higher than the average 4 % across all patient models. Detailed analysis of flow through the CoW as described in [16] indicates that R.ICA flow rate is directed towards the anterior of the CoW which allows for more particles to be influenced and routed by the high AcoA flowrate, which is directed from the right to left hemisphere. This potentially indicates that the AcoA is facilitating emboli from the right to left hemisphere, and preventing movement of emboli from left to right. These dynamics are clearly visible in the embolus trajectory animation video provided in Supplementary Material.

**Figure 6:**
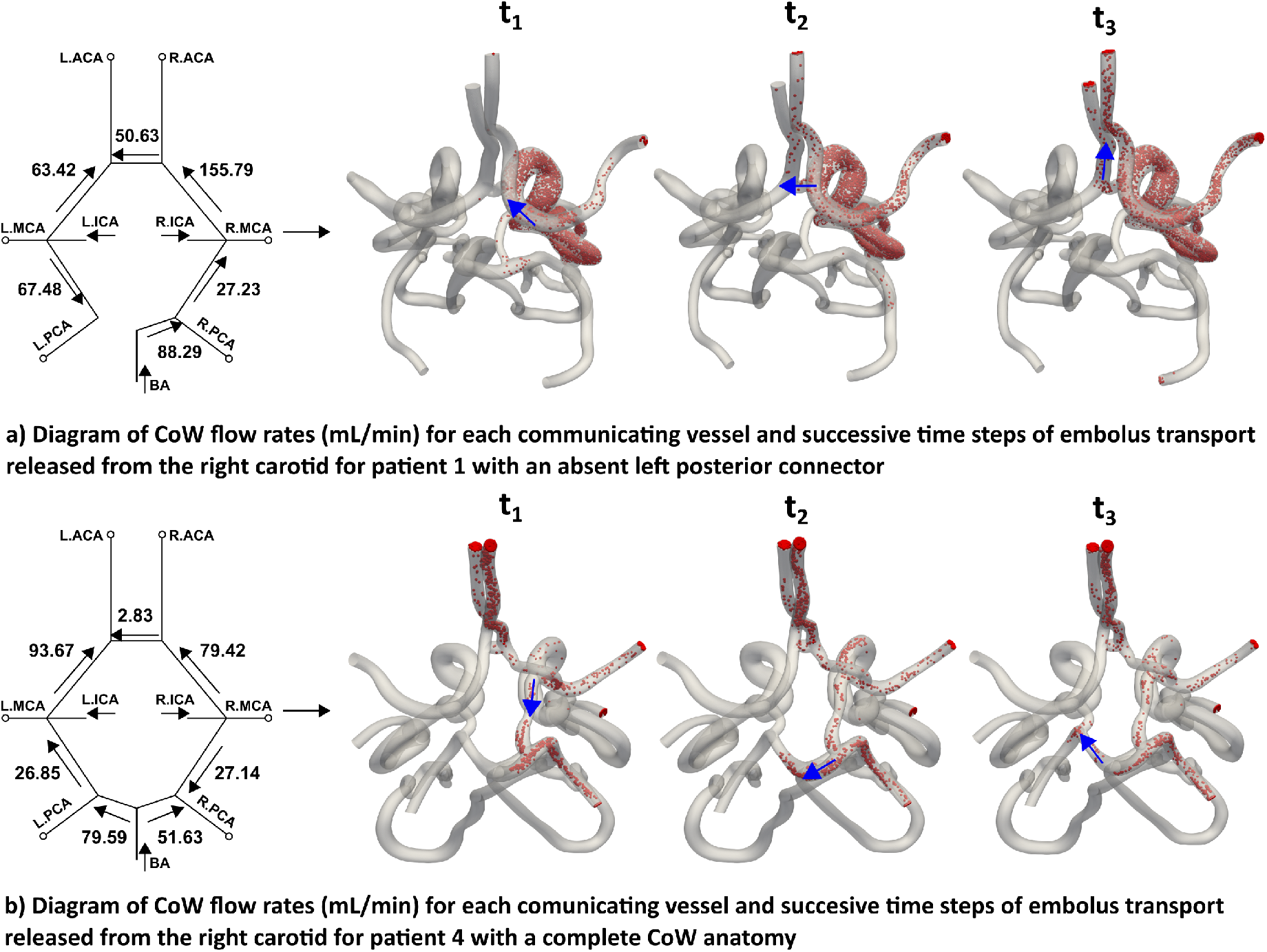
Two case studies examining how collateral proximal flow of the CoW affects the ability for contralateral movement. (a) Patient 1 with an absent left posterior connector exhibits how the collateral flow rate allows for contralateral movement from the right to left hemisphere through the AcoA. The AcoA has a high right to left flow rate that is able to route emboli into it and to the left hemipshere. (b) Patient 4 with a complete CoW anatomy completes contralateral movement thorugh the posterior region. The R.Comm flow rate directed towards the posterior routes emboli into the basilar artery and allows for travel into the left hemisphere due to the L.P1 flow rate directed towards the L.PCA.

Two additional observations are noted here based on detailed embolus trajectory data from our *in silico* experiments. First, while the AcoA was a key segment that enabled embolus transport contralaterally, it was not the only segment doing so. In Figure 6, panel b., we present another case of Patient 4 with a complete CoW network and emboli released from the right carotid site. In this case, emboli moved from the R.ICA into the R.Comm segment, then into the RP1 segment, and thereafter traveled over into the left PCA P1 segment. This embolus transport occurred over several cardiac cycles and is representative of the examples of non-intuitive embolus distribution dependent on flow-embolus interaction mechanisms in the CoW. Second, compared to systolic phase flow in the CoW vessels, diastolic flow exhibits significant retrograde and recirculating flow features. In several cases considered in this study, embolus interactions with these non-linear flow features led to complex non-intuitive trajectories. An example animation of such dynamics is provided in Supplementary Material, where 1 mm diameter thromboemboli move across Patient 3 CoW with a missing R.Comm segment. While initially systolic phase flow pushes the emboli into the ipsilateral A1 segment, the emboli interact with the retrogare diastolic flow and passes through the AcoA to move into the contralateral A1 and M1 segments. Both of these examples provide detailed evidence of highly non-intuitive embolus distribution patterns that: (a) differ from those expected from flow distributions alone; (b) differ from those expected from highly patent AcoA alone; and together they offer insights into mechanisms of contralateral transport that can be complicate etiology sourcing for carotid origin strokes.

The data and insights presented here have multiple broad clinical implications. The role of CoW vessels in collateral circulation in conjunction with the leptomeningeal collateral flow is acknowledged to be critical in acute ischemic stroke [33–36], even inspiring approaches towards collateral therapeutics [37]. However, less details are known about CoW collateral pathways being a route for emboli, and subsequent embolism sites in the brain - a potential contributing factor towards determining embolism etiology in strokes. Additionally, the observation that smaller emboli are more prone to contralateral movement and routing across CoW collateral pathways is of significance in small emboli pathological mechanisms that lead to microembolic showers [38], lacunar and watershed infarcts [39–41], and microstrokes leading to progressive vascular damage and dementia [42, 43]. Furthermore, understanding stroke risks from carotid stenosis has major implications on planning and outcomes of carotid revascularization therapy such as CAS or CAE [8]. Findings from this study are significant in advancing our understanding of peri- and post-operative stroke risks during revascularization procedures. A better understanding of when carotid lesions may lead to contralateral strokes could avoid false attributions of embolic sources and extensive or potentially harmful cardiac workups. It is relevant to note where such *in silico* models and analysis can find integration into direct patient cases. Currently, end-to-end computation time from standard-of-care images to embolus probabilities has an upper bound of 36-48 hours using reasonable state-of-the-art computational resources. Ongoing research is aiming to further reduce the compute time, for example, by accelerating image-segmentation using AI/ML based approaches. Hence, in their present form such *in silico* analyses would not be feasible within the rapid acute phase of standard-of-care which focuses on reperfusion and tissue recovery. However, post-acute phase secondary analysis of stroke etiology is potentially feasible wherein it would be akin to running an *in silico* test or lab and obtaining the work-up within a 36-48 hour (or less) time window. Finally, we have validated the one-way coupling embolus transport model using high resolution CFD [13], and validated the predictions and trends for embolism statistics against data reported in other clinical and imaging studies of cardiogenic embolus destinations [44, 45], and that of cardiogenic vs aortogenic embolus destinations [46] (demonstrated in prior work [14]). However, well controlled systematic comparison of the *in silico* framework against *in vitro* or benchtop models will further benefit the advancement of such modeling framework, which is an area of active investigation.

This study was based on several key assumptions. First, we assumed that the carotid sites did not have a visible severe stenosis. Presence of a severe stenosis will alter flow patterns through the carotid arteries into the CoW [47], which makes it difficult to isolate the effects of embolus-flow interactions at the CoW vessels alone. Furthermore, carotid arteries can be suspect to varying forms of embolization such as post-operative embolic showers after CEA [48] or endothelial trauma during catheterization, where stenosis severity will be low yet embolic phenomena may occur. These cases of non-symptomatic or mild stenosis can lead to otherwise unexpected or non-intuitive adverse cerebrovascular embolic events [49]. This aligns with the focus of this study on non-intuitive embolus transport in relation to flow distributions within the proximal collateral pathways as influenced by CoW anastomosis and embolus size. Additionally, the presence of higher-degree stenosis will require re-evaluation of hemodynamics as severe stenosis can alter hemodynamics within the heart-to-brain pathway. Incorporating these additional simulations within a parametric mechanistic study design as the one presented here would require a substantially more expensive computational effort that would be significantly beyond the scope of this paper. Nevertheless, the approach presented here can be extended in future studies to specifically account for stenosis of varying severity and its influence on cerebral embolic events. Along similar note, future efforts would also benefit from incorporating the simultaneous effects of contralateral carotid disease and vertebral artery disease to enable additional insights on embolic events from the cervical vessel sites. Second, we assume that the blood flow influences the emboli, but the emboli themselves do not significantly alter the flow (commonly referred to as one-way coupling). This is a reasonable assumption considering that each embolus is modelled individually as a single particle released independently of the other (that is, there is not a large concentration of thrombo-emboli suspended in blood to begin with). This assumption does limit our study to exclude cases where the embolus diameter approaches the sizes of the cerebral arteries themselves (that is, nearly occlusive emboli) [13]. To mitigate this, we focused on transport only to the M1/A1/P1 segments. However, the computational models can be further improved in future investigations if detailed models of flow disturbance around an individual embolus can be incorporated. Third, our physics-based embolus transport model assumes that the emboli are spherical particles. This is a widely-adopted modeling choice originating primarily from the fact that presently there is no generalized physics-model (such as Equation S5, for example) for arbitary non-spherical particle shapes, and derivation of a first-principles physics model of this nature will require significant additional theoretical and computational effort, which is beyond the scope of the study presented here. Additionally, embolus stretching, deformation, and deviation from sphericity is governed by the flow-induced forces, especially large gradations in shear forces as the embolus squeezes through a vessel. This effect is not dominant when we consider small-to-medium embolus sizes that do not approach the vessel diameters, which comprised the size regime we focused on this study. Finally, owing to the computational complexity of the overall set of simulations considered, we were able to consider 4 human subjects and 6 CoW anastomosis for each. In order to expand the interpretations further larger number of subjects need to be included, which is a substantial computational undertaking of its own beyond the scope of this current study. Regardless, the 144 experiments spanning over 1.7 million embolus samples, illustrates a rich set of evidence and mechanisms of contralateral embolus transport from carotid sites.

## 5 Concluding Remarks

We completed an investigation on how emboli originating from the carotid arteries may exhibit contralateral hemispheric movement when reaching the cerebral vessels. We completed a systematic *in silico* study comprising 144 embolus transport simulations by varying particle size, CoW anastomosis variation, and carotid side (left/right). The *in silico* framework enabled obtaining high resolution embolus transport data across patient-specific vascular anatomical models, with pulsatile hemodynamics - data that is otherwise challenging to obtain. 5 frequently observed CoW variations were modeled across 4 healthy patients in order to assess the effects of CoW anatomy on embolus transport influenced by underlying CoW collateral flow patterns. Contralateral or transhemispheric embolus movement was found to be significanty non-zero across the anatomical models regardless of carotid release. Smaller emboli demonstrated higher probability of traveling contralaterally in comparison to larger emboli. Proximal collateral flow rate of the CoW can promote contralateral movement in either hemispheric direction through the anterior or posterior communicating vessels. These findings can illuminate the mechanisms of contralateral movement that can contribute to sourcing non-decipherable emboli occlusion etiology.

## Supporting information

Supplementary Video: Case-Retrograde-Flow

Supplementary Video: Case-Fig-6b

Supplementary Video: Case-Fig-6a

Supplementary Vide: Embolus-CoW-Combined

## Data Availability

All quantitative embolus distribution data for this study have bee shared through the Open Science Framework as part of project titled Dataset: Transport and Distribution of Embolic Particles in Human Vasculature which can be accessed at: https://doi.org/10.17605/OSF.IO/CQKZT. All image-based modeling and computational hemodynamics simulations were conducted using the open source software package SimVascular. The image-based models (segmentations, pathlines), and computational hemodynamics data will be made available through the Vascular Model Repository associated with the SimVascular project, and can be accessed at: https://simvascular.github.io/. The sharing process with Vascular Model Repository is ongoing, and we expect sharing to be completed and data ready for access no later than 6 months from the publication of this article. For any data access issues, or for additional access to scripts and tools, please contact the corresponding author directly via email, or via the Contact page on the research teams official website: https://www.flowphysicslab.com/.

https://www.flowphysicslab.com/

## Conflicts of Interest

The Authors RR, ML, and DM have no conflicts of interest to report regarding this study.

## Acknowledgements

The Authors acknowledge funding support from National Institutes of Health Award: R21EB029736 for this study. This work utilized the Summit supercomputer, which is supported by the National Science Foundation (awards ACI-1532235 and ACI-1532236), the University of Colorado Boulder, and Colorado State University. The Summit supercomputer is a joint effort of the University of Colorado Boulder and Colorado State University. RR, and DM designed the study; RR conducted all simulations and data analysis; DM and ML interpreted simulation data; ML provided clinical insights; RR drafted the manuscript along with DM; RR, ML, and DM reviewed and edited the manuscript. All authors agreed to final version of the manuscript.

## Data, Materials and Code Disclosure Statement

All quantitative embolus distribution data for this study have bee shared through the Open Science Framework as part of project titled *“Dataset: Transport and Distribution of Embolic Particles in Human Vasculature”* which can be accessed at: https://doi.org/10.17605/OSF.IO/CQKZT. All image-based modeling and computational hemodynamics simulations were conducted using the open source software package Sim-Vascular. The image-based models (segmentations, pathlines), and computational hemodynamics data will be made available through the Vascular Model Repository associated with the SimVascular project, and can be accessed at: https://simvascular.github.io/. The sharing process with Vascular Model Repository is ongoing, and we expect sharing to be completed and data ready for access no later than 6 months from the publication of this article. For any data access issues, or for additional access to scripts and tools, please contact the corresponding author directly via email, or via the `Contact` page on the research teams official website: https://www.flowphysicslab.com/.

## Supplementary Material Information

### S1 Details on hemodynamics simulation

Here we present the foundational equations for the stabilized finite element hemodynamic simulations. Blood was modeled as a Newtonian fluid with a bulk density of 1.06 g/cc and viscosity of 4.0 cP. whose momentum and mass balance obeys the standard Navier-Stokes and continuity equations stated in Eq. S1 and S2 as stated below:

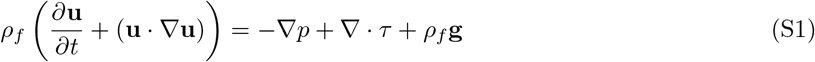

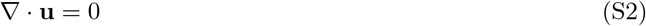

The term *τ* represents the viscous stresses in blood, which is modeled as per a linear Newtonian stress vs strain rate relation as follows:

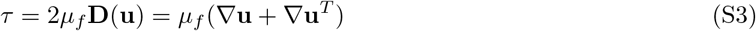

These equations are solved using matrix systems of equations obtained by decomposing a stabilized Petrov Galerkin finite element variational formulation of these equations over a mesh composed of linear tetrahedral elements. This variational formulation is stated in Eq. S4 as follows:

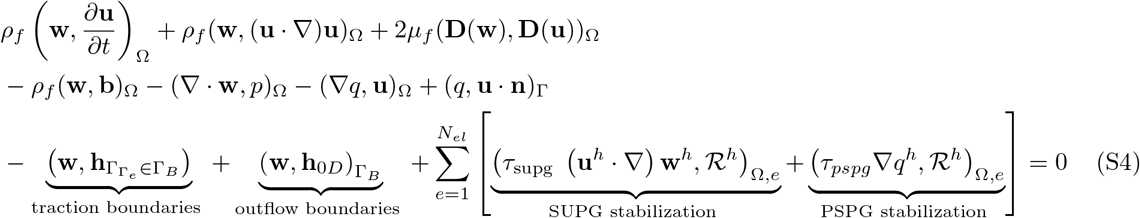

In these equations, **u**, p are blood flow velocity and pressure, **w**, q are the two test functions employed in the finite element solver, *μ*_*f*_ is the averaged blood viscosity, *ρ*_*f*_ is the averaged density of blood, **g** designates gravity, Ω denotes the computational domain of the arterial network, Γ and the subscripts of Γ denote the boundary faces of the computational domain, *ℛ*^*h*^ is the residual of the momentum equation, and *τ*_*supg/pspg*_ are the SUPG stabilization and PSPG stabilization factors used for convection and pressure stability respectively. **h**_0*D*_ represents the terms used to integrate the boundary conditions at the outlets and inlets of the arterial network; while 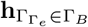 represents the other boundary conditions that may be necessary to run physiologically realistic flow simulations.

### S2 Details on embolus dynamics simulation

Embolic particle simulatons were conducted using a custom-modified version of the Maxey-Riley equation stated in Eq.S5 as below:

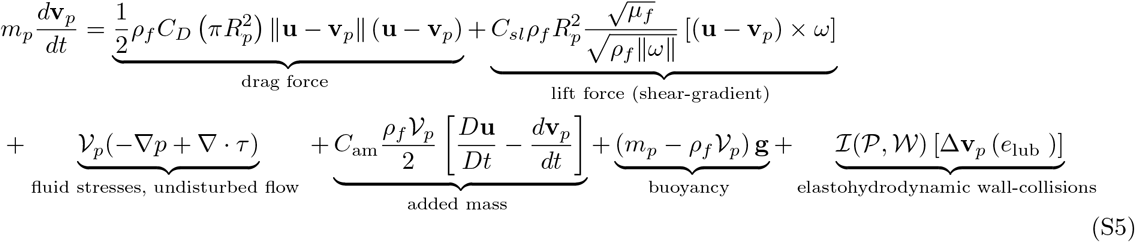

where m_*p*_ notes the particle mass, *𝒱*_*p*_ is the particle volume, R_*p*_ is the particle radius, *τ* denotes the stresses from undisturbed forces as defined in Section S1 and **v**_*p*_ is the translational particle velocity. C_*am*_ is the added mass coefficient, C_*D*_ is the drag coefficient, and C_*sl*_ denotes the shear-gradient lift force. *ω* = ∇ x **u** is the vortcity of the flow where the particle is located. ℐ (𝒫, 𝒲) is an indicator that has a value of 1 when a particle 𝒫 comes into contact with the artery wall, and the value of this function is 0 otherwise. Next, the contribution of the momentum changed created by the elastohydrodynamic lubrication and particle-wall collisions is denoted by the term [*δ***v**_*p*_(e_lub_)] where e_lub_ is the restitution coefficient.

### S3 Simple mathematical analysis of embolus inertia

Here we present a simple analysis of the dynamics of a spherical embolus particle when considering the drag forces and the buoyancy forces as stated in the Maxey-Riley based equation Eq. S5. Specifically, if we write the drag equation using a Stokes drag, and then divide throughout by the mass of a spherical particle, we can show that the equation reduced to the following form:

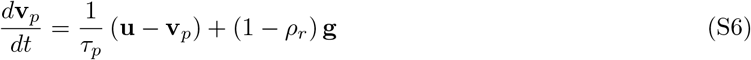

The term *τ*_*p*_ is essentially in the unit of time, and it refers to the time taken by a particle to respond to the background flow **u**. This is the momentum response time (MRT) as referred to in Section 4 in the main manuscript, and through simple algebra (not shown here) we can show that:

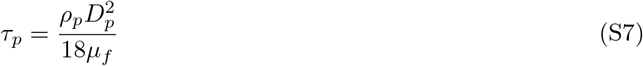

where *ρ*_*p*_ is the particle density, *D*_*p*_ is the diameter, and *μ*_*f*_ is the viscosity of background flow. For blood viscosity of 4.0 cP, and embolus density of around 1100.00 kg/m^3^, for a 1.0 mm diameter embolus this time is approximately 0.015 seconds. Considering that a standard cardiac cycle is of the order 0.8 seconds, this is approximately 1/40th of a cycle. While this is small, it has a finite impact on the trajectory because it will enable the particle to deviate from the background flow. A simple illustration of this can be shown by releasing a particle from a height *y* = *H* into a background flow such that it is uniform along the x-direction 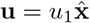. Assuming that the particle has a MRT of *τ*_*p*_ and density ratio *ρ*_*r*_, and that gravity acts along the y-direction (cross-stream from the flow), we can show that the particle velocity starting from rest, and the particle trajectory starting from *y* = *H* will be:

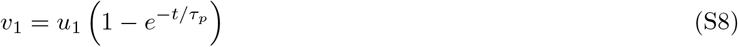

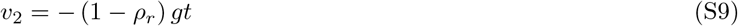

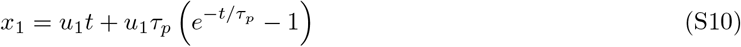

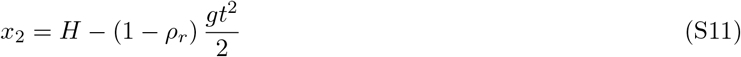

Simple trajectory analysis of this system is presented in Figure S1, and we can see that even at small values of MRT and for varying *ρ*_*r*_ the trajectories of the particles can differ from massless fluid parcels (MRT *≈* 0).

**Figure S1:**
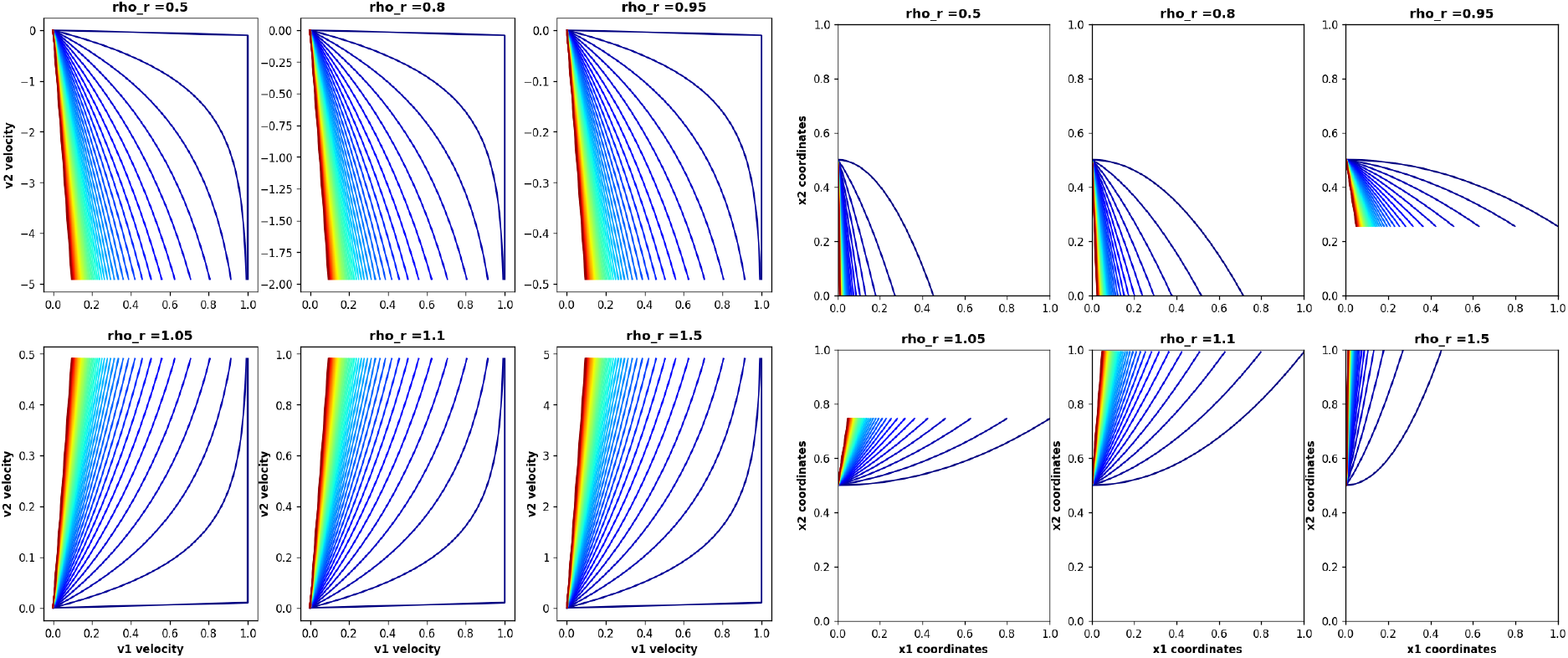
Sample trajectory and velocity data illustration based on the simplified analysis presented in Eq. S8-S11. The red indicates higher MRT, and blue indicates lower MRT.

### S4 Animation of embolus dynamics through the Circle of Willis

We describe the simulation animations that have been provided as additional illustration of the simulation data to support discussions presented in the main manuscript. Animation titled: ***Video-Embolus-CoW-Combined*.*mp4*** shows the thromboembolic particle transport dynamics for all 6 CoW variations for a single patient. This animation illustrates how emboli distribute into the CoW with emboli entering from the basilar artery and both internal carotids and distributing into the 6 CoW outlets. Left carotid emboli are colored blue and right carotid emboli are colored red. Contralateral movement can be seen clearly through the variations containing the AcoA with emboli crossing into the opposite hemisphere relative to the carotid release. Animation titled: ***Video-Supplementing-Case-Fig6a*.*mp4*** shows the data for Figure 6, panel a. We can observe closely the contralateral movement occurring through the AcoA segment, where emboli released from the right carotid travel into the CoW with the majority of emboli distributing ipsilaterally, and a significant proportion of emboli moving into the left hemipshere into the A1 segment through the AcoA. Animation titled: ***Video-Supplementing-Case-Fig6b*.*mp4*** shows the data for Figure 6, panel b. This illustrates the contralateral or transhemispheric movement of emboli through posterior vessel segments, showing that AcoA is not the only vessel that can play a role in such non-intuitive embolus distribution patterns. Lastly, animation titled: ***Video-Supplementing-Case-Retrograde-Flow*.*mp4*** demonstrates the scenario where embolus interaction with non-linear retrograde flow patterns in the CoW can lead to the non-intuitive distribution patterns for embolic particles in the brain. Retrograde movement of flow causes emboli to passively move into the AcoA and push into the opposite hemisphere during the next systole. This mechanism of fluid-particle interaction shows a less decipherable method of contralateral movement wherein average CoW vessel flow rates cannot directly inform due to it occurring during diastole.

## Notes

### Competing Interest Statement

The authors have declared no competing interest.

### Funding Statement

This study was funded by National Institutes of Health Award: R21EB029736. This work utilized the Summit supercomputer, which is supported by the National Science Foundation (awards ACI-1532235 and ACI-1532236), the University of Colorado Boulder, and Colorado State University.

### Author Declarations

The study used only imaging data for secondary retrospective analysis from a prior published work.

### Summary of Updates

We have made a change in title because the study did not actually consider diseased carotid artery sites with severe stenosis. We have included some more of our data for sharing, and embedded in the manuscript a link for shared data. Some additional clarifications about the image-data, and limitations of the computational model have been included.

